# RSV and all-cause lower respiratory infection burden among infants in remote British Columbia: Retrospective population-based birth cohort study

**DOI:** 10.64898/2026.01.09.26343745

**Authors:** Allison Watts, Marina Viñeta Páramo, Taylor Jamieson-Datzkiw, Jeffrey N. Bone, Léana Lavoie, Laura Arbour, Alfonso Solimano, Manish Sadarangani, Hind Sbihi, David Goldfarb, Pascal M Lavoie

**Author notes:** Corresponding author: Pascal M. Lavoie, MDCM, PhD, Department of Pediatrics University of British Columbia, British Columbia Children’s Hospital Research Institute 950 W 28th Ave, Vancouver, BC V5Z 4H4, Canada.

## Abstract

**Background:** In 2024, the National Advisory Committee on Immunization recommended universal RSV immunization across Canada, prioritizing infants in remote communities. However, in the absence of population-based data, programs may not effectively narrow health gaps in remote communities.

**Methods:** Retrospective cohort study of all births in British Columbia (BC) from April 2013 to March 2024, followed for 1 year, using health administrative data. Main outcomes were hospitalizations for all-cause and RSV-lower respiratory tract infection (LRTI). Secondary outcomes were tertiary Pediatric Intensive Care Unit (PICU) admissions, length of stay, and air transport. Main exposures were community remoteness and social determinants of health. Incidence rates and incidence rate ratios (IRR) adjusted for sex, prematurity, and chronic conditions were estimated using Poisson generalized estimating equations.

**Results:** Among 472,623 infants, those living in remote communities (N=3636) had higher hospitalization risk for all-cause (IRR: 2.91, 95%CI 2.02-3.65) and RSV-LRTIs (IRR: 1.60, 95%CI 1.17-2.19) compared to metropolitan areas. Length of stay and PICU admission rates were similar by region. Almost half (48.8%) of children from remote areas hospitalized for all-cause LRTIs required air evacuation. Infants from remote communities remained at higher risk for all-cause (aIRR 2.84, 95% CI 2.22–3.63) and RSV-LRTI (aIRR 1.56, 95% CI 1.15–2.12) hospitalizations after adjusting for covariates.

**Interpretation:** Infants in remote communities experienced a disproportionately high RSV-LRTI burden, supporting prioritized RSV interventions in these regions. The residual risk after accounting for known factors highlights the need to investigate additional drivers of vulnerability in remote areas.

## INTRODUCTION

Respiratory syncytial virus (RSV) is the leading cause of lower respiratory tract infection (LRTI) and hospitalization in children under one year of age^1–3^. In 2024, the National Advisory Committee on Immunization (NACI) recommended that provinces and territories build toward universal RSV immunization programs for all infants. NACI also advised prioritizing infants whose transportation for severe RSV disease treatment is complex and/or whose risk intersects with established social and structural health determinants such as those experienced by some Indigenous communities^4,5^.

Infants in remote areas often require medical air transport for hospitalizations, creating substantial strain on families and health systems. National surveillance data primarily capture admissions to tertiary pediatric centers^6^, and therefore underestimate the true burden of RSV in remote regions. A study from Nunavut showed that most RSV-related hospitalizations among infants from remote communities occur in regional facilities^7^. However, prior hospital-based studies have not provided population-level estimates of RSV burden or risk factors across gradients of remoteness. Such analyses are essential for equitable policy decisions and for directing resources where they are most needed.

Although socioeconomic factors contributing to RSV burden have been explored,^8–11^ reasons for persistent disparities remain poorly understood. These factors shape healthcare access, uptake of preventive interventions and overall program impact. Identifying community-level factors is critical to achieving equitable immunization coverage and maximizing the benefit of RSV prevention strategies.

The objectives of this study were to: (1) quantify the burden of all-cause and RSV-LRTI hospitalizations among infants living in remote communities in British Columbia (BC) and (2) examine individual and community-level risk factors, including social determinants of health.

## METHODS

### Study design, participants and setting

This is a retrospective birth cohort study including all infants born in BC over an 11-year period (April 2013 to March 2024) followed to one year of age.

BC has a population of 5.7 million in 2024 and an average of approximately 42,000 infants per year were born over the past 5 years. Children were identified through enrollment in the provincial public health services plan within their first 30 days after birth (>98% of all live births in BC). Data were obtained from the BCC19C administrative dataset linking the following BC Ministry of Health datasets: Hospital Discharge Abstract Database (DAD), Medical Services Plan (MSP) Client Roster, pharmacy records (PharmaNet), Perinatal Data Registry (PDR), and the 2021 Statistics Canada Census. Children were excluded if their registration date in the provincial health service plan could not be determined.

The study was conducted using de-identified data as part of a program evaluation initiative; therefore, ethics review was waived and they study was approved by the Ministry of Health of BC.

Outcome measures

The primary outcomes were hospital admissions for all-cause LRTIs or RSV-LRTIs. For RSV-LRTIs, based on ICD-10 codes (Supplementary Table 1), we previously established that this outcome had 81% sensitivity and 98% specificity capturing hospitalizations due to RSV in children under 2 years of age in BC, when compared against viral testing data^12^. Specificity for RSV hospitalizations remained high for all remoteness categories, with lower sensitivity in rural and remote communities (Supplementary Table 2). Hospitalizations within 30 days of one another were considered as the same episode of infection.

Secondary outcomes were all-cause LRTI or RSV-LRTI tertiary Pediatric Intensive Care Unit (PICU) admissions; length of hospitalization (in days); length of PICU stay (in days); and use of medical transportation by air.

### Exposures

The main exposures were remoteness of residency location, assigned for each infant as one of four categories (Remote, Rural, Urban, and Metropolitan) and social determinants of health.

Remoteness was defined by place of residence at birth (or the earliest residential information available), using the BC Ministry of Health classification according to population size and a remoteness index attributed to each Community Health Service Area (CHSA) in BC.^13^ CHSA boundaries changed between 2018 and 2022. Remoteness categories from 2018 were used for babies born prior to 2020, all others used the 2022 boundaries. The current geographical distribution of CHSA by remoteness categories is illustrated in Figure 1.

**Figure 1.**
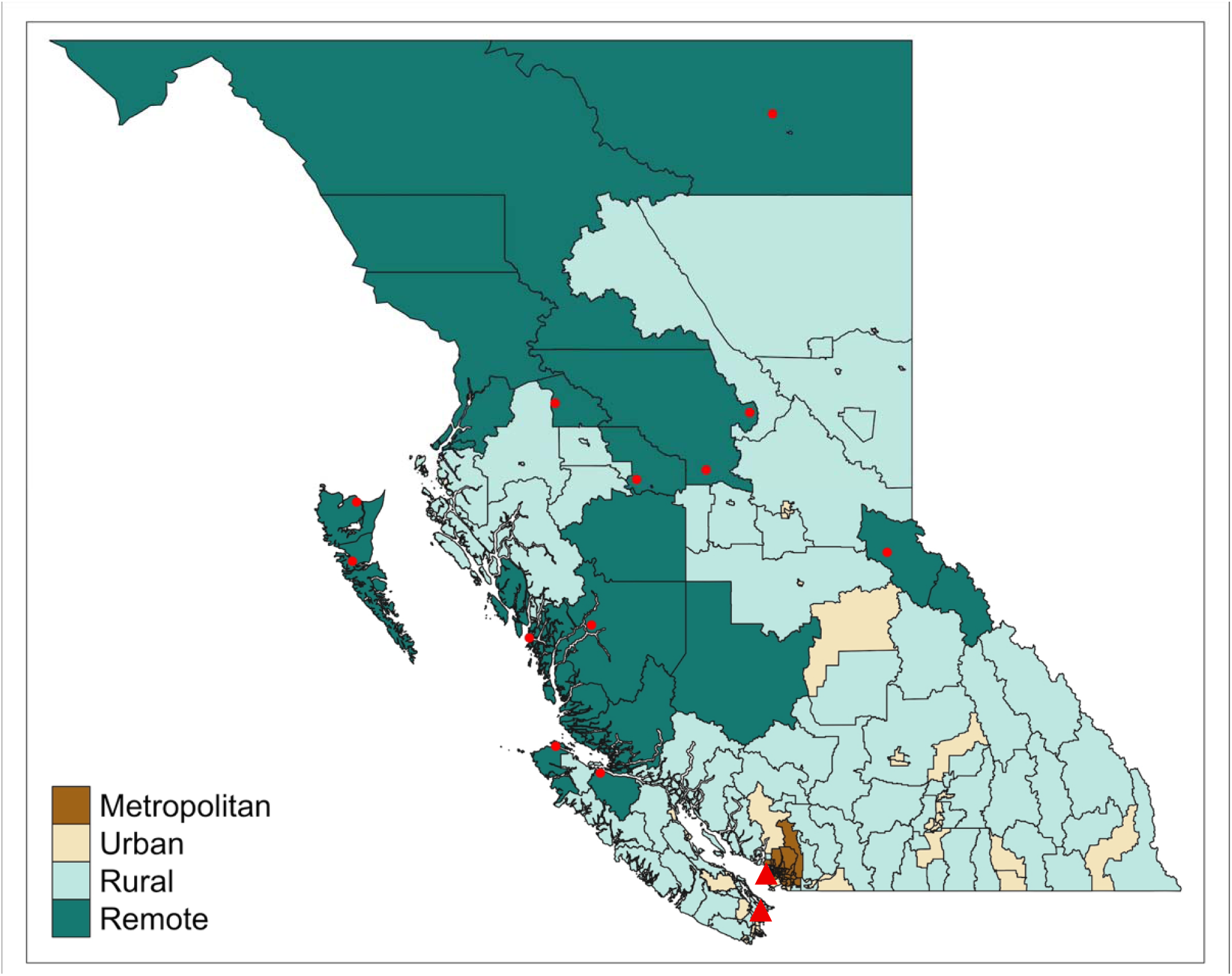
Remote, rural, urban and metropolitan communities in British Columbia, Canada Red dots denote hospitals in remote communities, red triangles denote tertiary care pediatric intensive care hospitals^13^

Eligibility for income assistance was based registration for Plan C in the provincial database of pharmacy records, PharmaNet. Plan C provides coverage for those receiving income assistance through the BC Ministry of Social Development and Poverty Reduction and children in the care of the Ministry of Children and Family Development.^14^ It was used as a measure of an infants’ family income status. All babies with a Plan C registration in their first two years of life, or whose mothers were registered with Plan C within 2 years of their birth were coded as being eligible for income assistance. All others were coded as not eligible.

British Columbia Index of Multiple Deprivation (BCIMD) are measures of deprivation at the level of the CHSA using the 2022 CHSA boundaries and the 2021 Census and Statistics Canada’s Canadian Index of Multiple Deprivation methodology to create four distinct dimensions of deprivation: Situational Vulnerability (proportion of population aged 25-62 without a high-school diploma, of dwellings needing major repairs, that is low income, and of single parent families); Economic Dependency (proportion not in labor force, aged 65 and older, unemployment ratio, dependency ratio: those aged 0-14 and 65+ divided by those aged 15-64); Ethno-cultural Composition (proportion of population that are visible minorities, foreign-born, have no knowledge of official languages, recent immigrants in the last 5 years); and Residential Instability (proportion of dwellings that are apartment buildings, persons living alone, dwellings that are not owned, moved within the past five years)^15–17^. All variables were categorized so that the fifth quintile represents the group with the highest level of deprivation. The BCIMD suppressed the results from one CHSA because of missing information (n=236, 0.05%).

Covariates: Sex, prematurity (categorical: <28 and 28+0 to 36+6, and ≥37 weeks of gestation), and presence of a chronic medical condition^12^.

### Statistical analyses

The incidence rate (IR) (number of events divided by the follow-up in person-years) of all cause LRTI and RSV-LRTI hospitalizations with 95% confidence intervals (CI) were estimated across community remoteness and indicators of social determinants of health (income assistance and community-level BCIMD dimensions). In addition, the IR of PICU admissions was estimated across community remoteness levels. Among those hospitalized, median days and total bed-days and IQR for hospital and PICU admissions, and proportion requiring medical transportation use were described for each category of community remoteness. Infants who died or moved out of province within 12 months of age were censored at their last follow-up date (n=1137, 0.2%).

Generalized estimating equations (GEE) were used to estimate the risk of all-cause LRTI and RSV-LRTI hospitalizations, on average, for children living in remote communities compared to metropolitan communities and to see if this average risk was independent of other known risk factors that may be present in remote communities such as differences in clinical risk factors or social disadvantage. GEE models included a Poisson distribution, log time in person-years as the offset and clustering of individuals by community (CHSA).^18^ Metropolitan communities were used as the reference category as they were the largest group, and had the lowest incidence rates. Models were run for each variable separately and then adjusted models were used to look at the effect of remoteness on hospitalization independent of each social determinant of health variable with additional adjustment for covariates: sex, being born prematurely, and presence of a chronic medical condition.

To better understand the role of specific social determinants of health within remote communities, we plotted the IR of hospitalization in each community type by each social determinant of health indicator and created stratified models showing the incidence rate ratio (IRR) of hospitalization for each social determinant of health indicator (adjusted for individual factors) among children living in each community type. In all models, results are presented as IRR or adjusted IRR (aIRR) and 95% CI. Models were run separately for all-cause LRTI and RSV-LRTI outcomes.

## RESULTS

### Cohort characteristics

The cohort comprised 472,623 infants, with the distribution across community remoteness categories summarized in Table 1. Sex distribution, rates of prematurity, and the prevalence with chronic medical conditions were similar across community types (Table 1; Supplementary Table 3).

**Table 1.** Baseline characteristics of infants by community remoteness (n=472,623).

|  | **Remote** | **Rural** | **Urban** | **Metropolitan** |
| --- | --- | --- | --- | --- |
| N (%) | 3636 (0.8%) | 63,762 (13.5%), | 164,756 (34.9%) | 240,469 (50.9%) |
| Sex, male, n (%) | 1830 (50.3%) | 32,814 (51.5%) | 84,499 (51.3%) | 123,816 (51.5%) |
| Prematurity, n (%) |  |  |  |  |
| >37 weeks | 3268 (89.9%) | 59,026 (92.6%) | 151,613 (92.0%) | 222,664 (92.6%) |
| 28-37 weeks | 356 (9.8%) | 4574 (7.2%) | 12,599 (7.7%) | 17,036 (7.1%) |
| <28 weeks | 11 (0.3%) | 162 (0.3%) | 545 (0.3%) | 769(0.3%) |
| At least one chronic medical condition, n (%) | 198 (5.5%) | 3257 (5.1%) | 9626 (5.8%) | 15,233 (6.3%) |

### Burden of all-cause and RSV-LRTI by community remoteness

Overall, 7,186 (1.52%) and 3,901 (0.83%) infants were hospitalized at least once for all-cause or RSV-LRTIs in the first year of life, respectively (Table 2). Hospitalization rates significantly increased among infants living in more remote communities for both all-cause and RSV-LRTI. For instance, hospitalization rates ranged from 35.6 [95%CI 29.8-42.4] for all-cause LRTI for infants living in remote communities, compared with 12.2 [11.8-12.7] for those living in metropolitan areas (Figure 2). For RSV- LRTI, the hospitalization rate was 11.3 [95%CI 8.1-15.4] for infants in remote communities compared to 7.1 [95%CI 6.7-7.4] for those in metropolitan communities.

**Figure 2.**
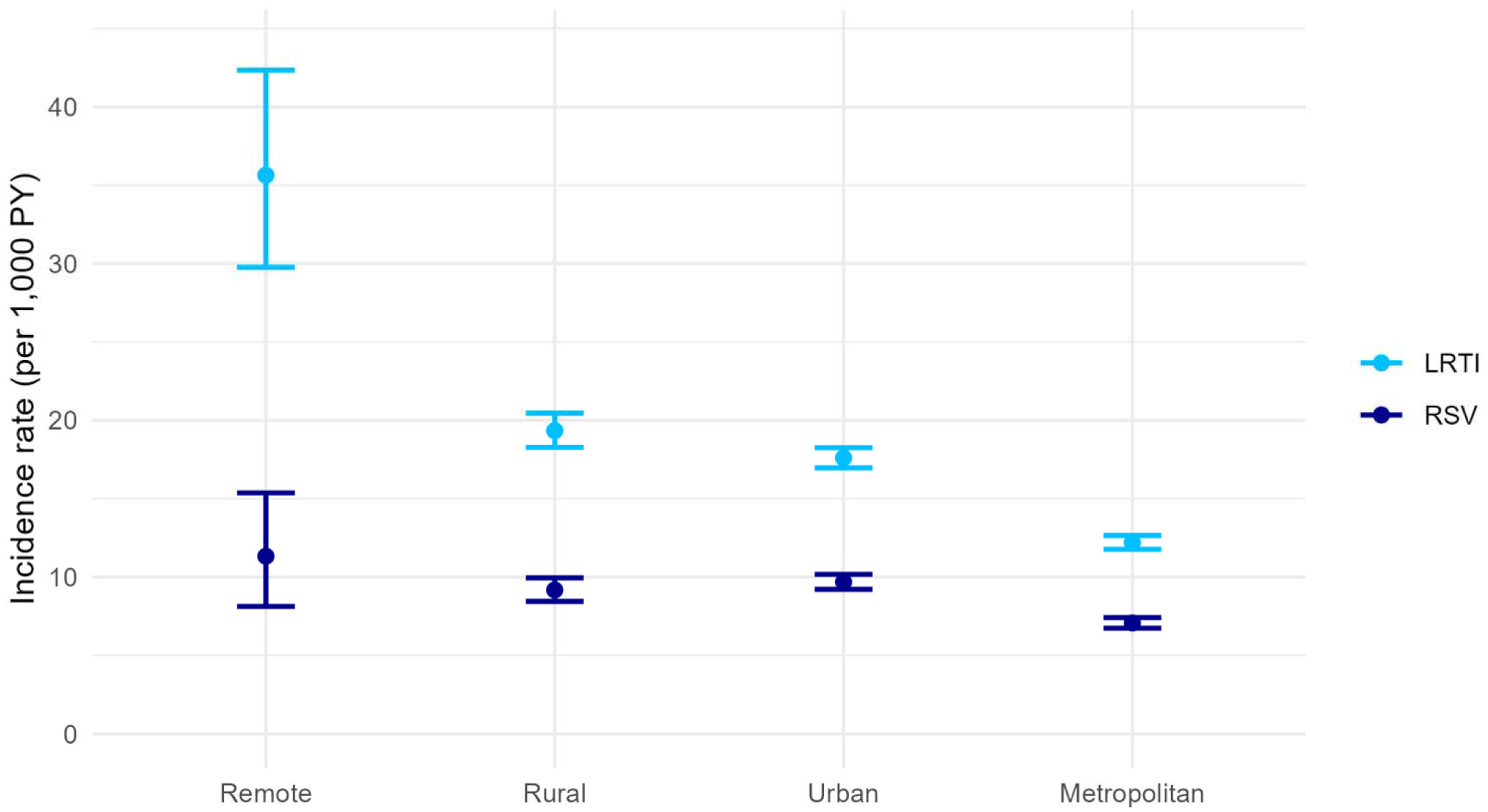
Incidence rate of all-cause LRTI and RSV-LRTI hospital admissions by community type in British Columbia, infants < 1 years old, 2013-2024 (with 95%CI).

**Table 2.**
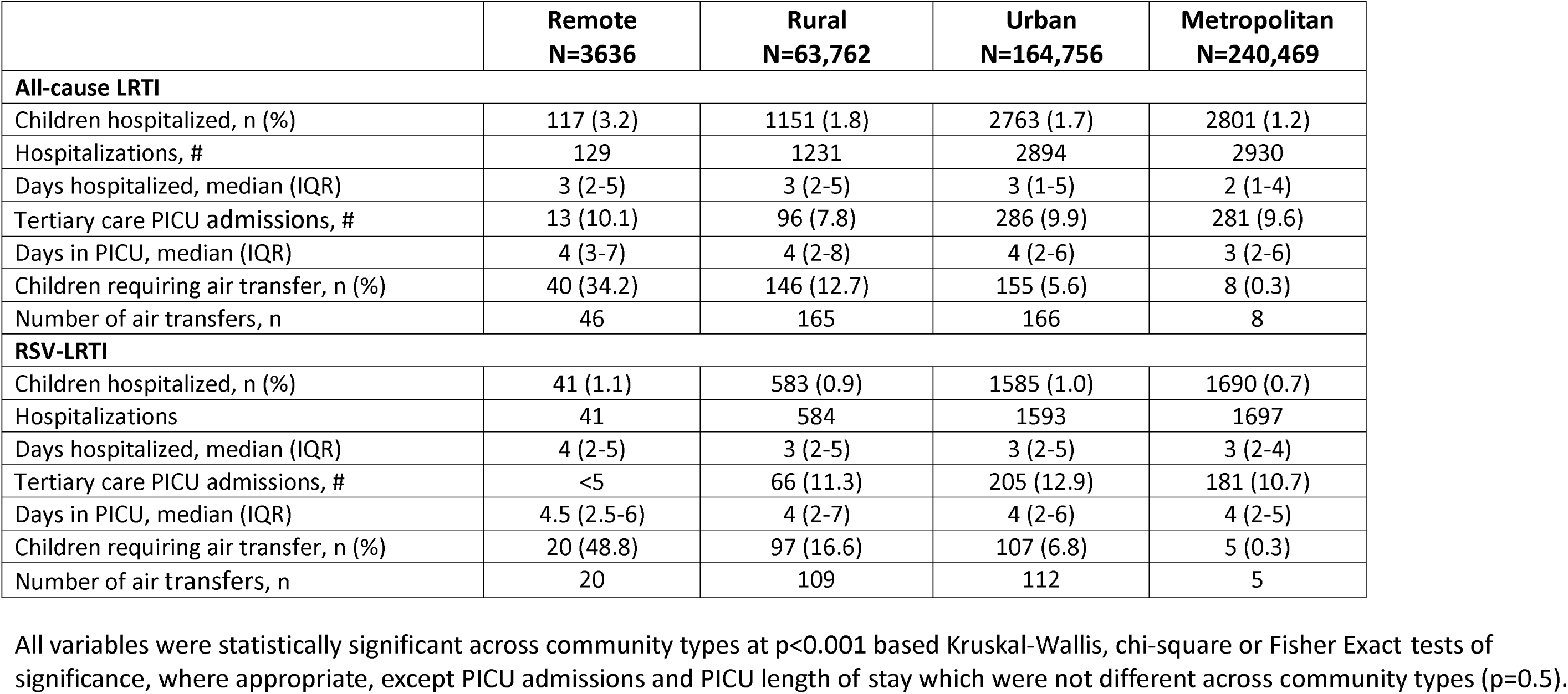
All-cause LRTI and RSV-LRTI hospitalization outcomes among infants by community remoteness (n=472,623). All variables were statistically significant across community types at p<0.001 based Kruskal-Wallis, chi-square or Fisher Exact tests of significance, where appropriate, except PICU admissions and PICU length of stay which were not different across community types (p=0.5).

Length of hospital stay, the proportion of infants requiring tertiary PICU stay and their length of PICU stay did not differ across community groups, suggesting comparable degree of infection severity (Table 2). However, in sharp contrast one third (34.2%) and almost half (48.8%) of infants in remote communities hospitalized for all-cause and RSV-LRTI required air transport, with proportions that are much higher among infants living in more remote, compared to metropolitan regions (Table 2).

### Individual and community level social and economic characteristics

Marked differences in indicators of social determinants of health were observed across community types, with remote communities showing substantially greater situational vulnerability, characterized by limited access to services or higher unemployment, than any other community type (Table 3).

**Table 3.** Social determinants of health by community remoteness categories (n=472,623). Economic dependency, situational vulnerability, residential instability and ethno-cultural composition measured at the community level (community health services area) and quintiles are from least deprived (Q1) to most deprived (Q5).

|  | **Remote** | **Rural** | **Urban** | **Metropolitan** |
| --- | --- | --- | --- | --- |
| N (%) | 3636 (0.8%) | 63,762 (13.5%), | 164,756 (34.9%) | 240,469 (50.9%) |
| Eligibility for income assistance, n (%) | 235 (6.5%) | 4854 (7.6%) | 14,239 (8.6%) | 11,225 (4.7%) |
| Situational vulnerability, n (%) |  |  |  |  |
| Q1 | 0 | 8719 (13.7%) | 35,381 (21.5%) | 41,909 (17.4%) |
| Q2 | 0 | 18,455 (28.9%) | 30,089 (18.3%) | 59,876 (24.9%) |
| Q3 | 175 (5.2%) | 16,024 (25.1%) | 35,431 (21.5%) | 50,184 (20.9%) |
| Q4 | 294 (8.7%) | 9,273 (14.5%) | 27,541 (16.7%) | 67,431 (28.0%) |
| Q5 | 2931 (86.2%) | 11,291 (17.7%) | 36,315 (22.0%) | 21,069 (8.8%) |
| Economic dependency, n (%) |  |  |  |  |
| Q1 | 513 (15.1%) | 10,848 (17.0%) | 33,986 (20.6%) | 84,567 (35.2%) |
| Q2 | 0 | 12,839 (20.1%) | 45,224 (27.5%) | 80,290 (33.4%) |
| Q3 | 821 (0.8%) | 11,483 (18.0%) | 37,101 (22.5%) | 50,267 (20.9%) |
| Q4 | 1529 (45.0%) | 10,646 (16.7%) | 31,583 (12.2%) | 23,185 (9.6%) |
| Q5 | 537 (15.8%) | 17,946 (28.2%) | 16,862 (10.2%) | 2160 (0.9%) |
| Residential instability, n (%) |  |  |  |  |
| Q1 | 1234 (36.3%) | 22,139 (34.7%) | 21,283 (12.9%) | 37,096 (15.4%) |
| Q2 | 376 (11.1%) | 19,828 (31.1%) | 25,115 (15.2%) | 27,126 (11.3%) |
| Q3 | 775 (22.8%) | 12,318 (19.3%) | 32,803 (19.9%) | 42,984 (17.9%) |
| Q4 | 939 (27.6%) | 8369 (13.1%) | 52,857 (32.1%) | 29,809 (12.4%) |
| Q5 | 76 (2.2%) | 1108 (1.7%) | 32,698 (19.9%) | 103,454 (43.0%) |
| Ethno-cultural composition, n (%) |  |  |  |  |
| Q1 | 903 (26.6%) | 22,778 (35.7%) | 12,387 (7.5%) | 0 |
| Q2 | 654 (19.2%) | 20,873 (32.7%) | 47,601 (28.9%) | 0 |
| Q3 | 1243 (36.6%) | 10,646 (16.7%) | 54,542 (33.1%) | 7926 (3.3%) |
| Q4 | 600 (17.6%) | 5895 (9.3%) | 42,661 (25.9%) | 74,816 (31.1%) |
| Q5 | 0 | 3570 (5.6%) | 7565 (4.6%) | 157,727 (65.6%) |

Greater economic dependency, reflecting a greater proportion of residents not participating in the labour force relative to those employed, was observed outside metropolitan communities.

Conversely, remote populations had lower residential instability, reflecting less frequent movement of residents into or out of communities. Ethno-cultural diversity was also markedly more characteristic of metropolitan communities, where it is less common in remote or rural communities. The proportion of infants whose families qualified for income assistance was similar across community types (Table 3).

### Associations between social determinants of health and LRTI risk

Specific social determinants of health differentially affected LRTI hospitalization risk between community types. This is shown graphically in Figure 3, with exploratory statistical analyses for each indicator presented in Supplementary Table 5. For instance, situational vulnerability was a strong predictor of hospitalization risk in remote communities after adjustment for sex, prematurity and chronic medical conditions for RSV-LRTI (aIRR 1.51, 95% CI 1.20–1.90) and all-cause LRTI (aIRR 2.09, 95% CI 0.97–4.51), (Figure 3; Supplementary Table 5). Eligibility for income assistance was associated with higher hospitalization risk for all-cause or RSV-LRTI among rural, urban and metropolitan communities, where residential instability was associated with higher hospitalization risk for all-cause and RSV-LRTI in population from metropolitan communities.

**Figure 3.**
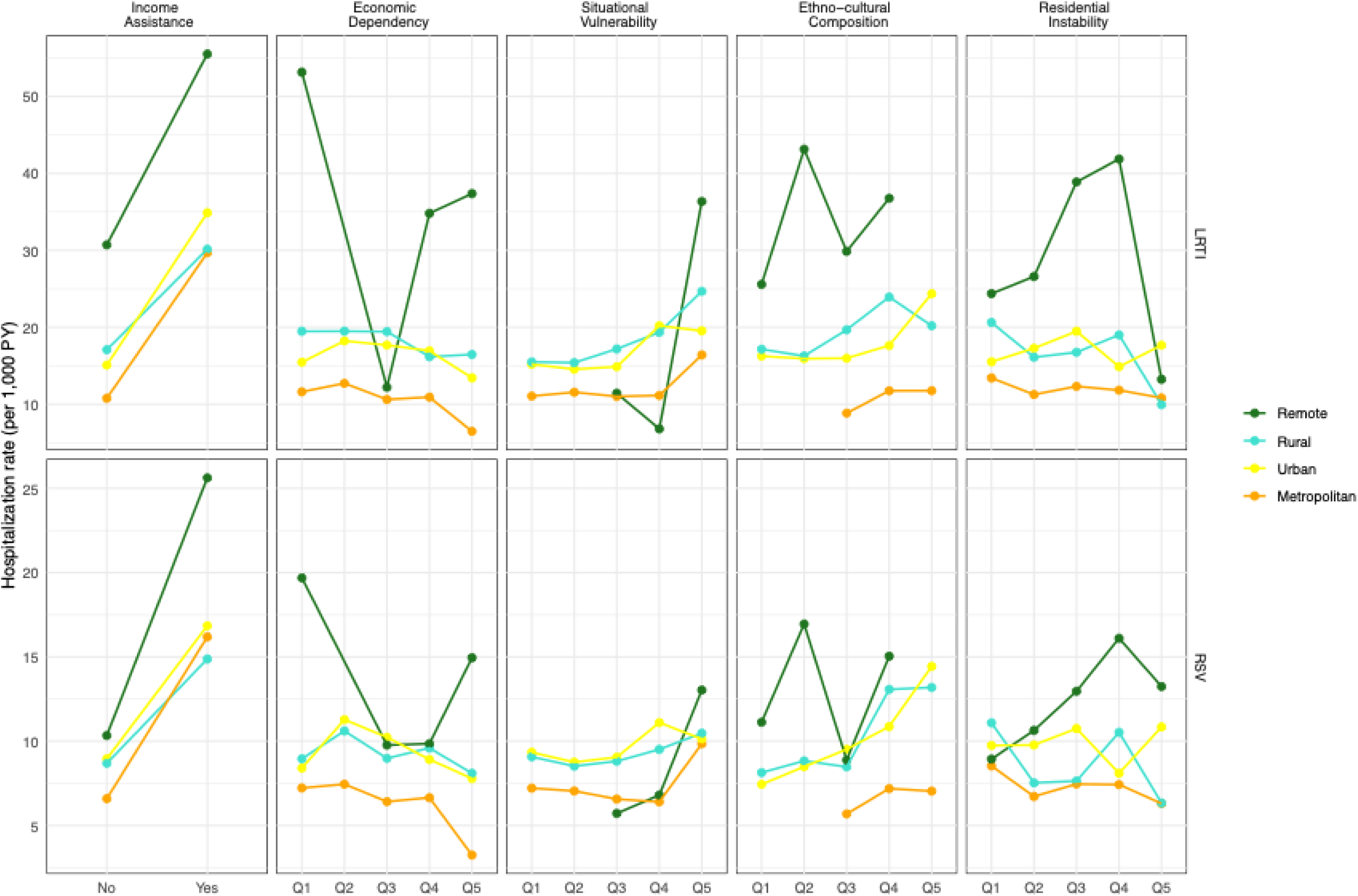
Incidence of all-cause LRTI and RSV-LRTI hospitalizations by community type and social determinants of health, presented as quintiles from least deprived (Q1) to most deprived (Q5).

In multivariable models adjusting risk of all-cause and RSV-LRTI for social determinants of health by community remoteness, eligibility for income assistance, greater community situational vulnerability and lower community ethno-cultural composition were significantly associated with an increased risk of all-cause LRTI hospitalization, across communities (Supplementary Table 4). Similar associations were observed for RSV-LRTI, except that the association of situational vulnerability was attenuated and lower residential instability was associated with an increased risk of hospitalization. Altogether, these analyses highlighted the significant influence of social determinants of health indicators on the risk of hospitalization in infants with all-cause or RSV-LRTI.

Importantly, infants from remote communities remained at significantly higher risk of hospitalization compared with those from metropolitan areas (aIRR 2.84, 95% CI 2.22–3.63 for all-cause LRTI; aIRR 1.56, 95% CI 1.15-2.12 for RSV-LRTI) after adjustment of individual factors and social determinants of health (Table 4).

**Table 4.** Unadjusted and adjusted incidence rate ratios of all-cause and RSV-LRTI hospitalizations by community remoteness. Poisson regression accounting for clustering within community health service areas; IRR=Incidence rate ratio; ^a^adjusted models show the association between community remoteness and hospitalization risk independent of sex, prematurity, chronic medical conditions and eligibility for income assistance; full models shown in supplementary Table 4

|  | Unadjusted | Adjusted<sup>a</sup> |
| --- | --- | --- |
|  | Hospitalization for all-cause LRTI, IRR (95% CI) |  |
| Remote | 2.91 (2.24-3.79) | 2.84 (2.22-3.63) |
| Rural | 1.59 (1.38-1.83) | 1.55 (1.36-1.78) |
| Urban | 1.44 (1.28-1.63) | 1.37 (1.23-1.53) |
| Metropolitan | Reference | Reference |
|  | Hospitalization for RSV-LRTI, IRR (95% CI) |  |
| Remote | 1.60 (1.17-2.19) | 1.56 (1.15-2.12) |
| Rural | 1.30 (1.13-1.50) | 1.27 (1.11-1.46) |
| Urban | 1.37 (1.22-1.55) | 1.32 (1.18-1.48) |
| Metropolitan | Reference | Reference |

## DISCUSSION

In this population-based study, we found that although infants from remote communities accounted for fewer than 1% of all births in BC, they experienced a disproportionately high risk of hospitalizations for both all-cause and RSV-LRTIs. As expected, indicators of disease severity, such as the proportion requiring admission to a tertiary pediatric intensive care unit and length of stay, were similar across communities; however, infants from remote regions required substantially more frequent air transport for hospital care. This study provides province-wide, population-level evidence quantifying disparities in LRTI hospitalization outcomes and their associations with social determinants of health across community types, supporting NACI’s recommendation to prioritize remote populations as provinces work toward implementing new RSV prevention programs. These data highlight the need for region-specific economic evaluations, and for targeting resources to maximize impact in these communities. To our knowledge, this is the first population-level study focused on remote communities, offering critical data to guide the implementation and evaluation of RSV immunization programs.

While several provinces have moved toward universal RSV immunization, early implementation appears to have overlooked the unique challenges faced by remote communities. Equity in program delivery cannot be assumed; it must be deliberately built into design and evaluation. Data on uptake, timeliness, and access in remote regions should be systematically collected, and specific measures, such as community-led outreach or dedicated distribution pathways, should be implemented to ensure that the infants at highest risk are not left behind.

A notable strength of this study is that it captured virtually all births in an integrated health jurisdiction, with validated hospitalization outcomes and social determinant of health indicators. Inclusion of regional hospitals allowed us to capture admissions that would otherwise be missed by tertiary-care-based surveillance, providing a more complete picture of the burden in remote settings.

Community-level situational vulnerability, defined by indicators such as income, educational attainment, and the proportion of single-parent families, was linked to increased hospitalization risk in remote communities. This finding highlights the need for comprehensive public health strategies that extend beyond immunization, addressing upstream social determinants of health that contribute to the elevated burden of disease in these communities. Disparities persisted after adjustment for demographic and clinical factors as well as individual and community-level social determinants of health; therefore, these data support the need for immunization, but also for a broader approach including interventions to address structural health inequities in those communities.

Previous studies have attributed differences in LRTI hospitalization risk to socioeconomic conditions, structural health system factors, breastfeeding, low birth weight or other environmental factors, including indoor air quality, smoking, and inadequate plumbing^8–11,11,19–21^. Importantly, the persistence of elevated risk after adjusting for biological and social determinants of health in this study suggest that additional biological or contextual factors contribute to vulnerability in these populations.

Increased LRTI hospitalization risk has been noted in Canadian Inuit infants, or rural or remote Indigenous populations in Alaska and Australia ^7,8,10,11,19–21^. Canadian case studies have highlighted up to tenfold regional differences in RSV hospitalization rates among infants in remote Indigenous communities.^7,19^ U.S. studies have demonstrated that Indigenous infants experience substantially higher RSV hospitalization rates^22,23^. In BC, a carnitine palmitoyltransferase 1A (CPT1A) gene variant, disproportionately prevalent in Indigenous coastal populations, has been associated with increased susceptibility to severe respiratory illness and other infection^8,24^. This hypomorphic variant impairs lipid metabolism, and may affect susceptibility to infection^25^. Collectively, the findings warrant further investigations to understand determinants of vulnerability amendable beyond potentially stigmatizing socioeconomic factors. These findings highlight the need for strengthened culturally appropriate prevention strategies to equitably mitigate pediatric LRTI morbidity.

This study has limitations. Although LRTI hospitalizations were identified using ICD-10 codes, potential regional differences in testing or coding practices may influence case ascertainment. To mitigate this, we examined both RSV-specific and all-cause LRTI outcomes, which yielded similar patterns.

Additionally, these ICD-10 codes do not have perfect sensitivity and therefore misclassification may bias results. The way in which social determinants of health variables are related to each other and community remoteness is not straightforward (e.g., the direction of effects or whether on the causal pathway), thus different modeling approaches or different measures of social determinants of health or remoteness may lead to different associations than those presented here. Community-level social variables were drawn from 2021 census data and 2022 geographical boundaries and may not reflect temporal changes. Further, these community-level social variables do not apply directly to all individuals in an area and are subject to ecological biases when extrapolating estimates to the individual level.

In conclusion, infants in remote communities in BC experience substantially higher rates of hospitalization for RSV and other LRTIs, after accounting for measured clinical and social indicators. These findings underscore the need for targeted RSV prevention strategies, careful monitoring of immunoprophylaxis program impact, and broader policies addressing the structural and environmental determinants that perpetuate respiratory health inequities in remote populations.

## AUTHOR CONTRIBUTIONS

Concept and design: Watts, Viñeta Paramo, Jamieson-Datzkiw, PM Lavoie Acquisition, analysis, or interpretation of data: Watts, Viñeta Paramo, Jamieson-Datzkiw, L Lavoie, Bone, Arbour, Sbihi, Goldfarb, PM Lavoie Drafting of the manuscript: Watts, PM Lavoie Critical review of the manuscript for important intellectual content: Viñeta Paramo, Jamieson-Datzkiw, Bone, Solimano, Wiens, Arbour, Sbihi, Goldfarb Statistical analysis: Viñeta Paramo, Watts, Bone Obtained funding: PM Lavoie Administrative, technical, or material support: N/A Supervision: PM Lavoie Drs Watts and Lavoie had full access to all the data in the study and take responsibility for the integrity of the data and the accuracy of the data analysis.

## Data Availability

Requests for the data dictionary and/or statistical/analytic code may be directed to. Access to line-level, de-identified data is subject to approval by the Data Stewards and applicable data-sharing agreements and ethics approvals. These data may be requested for scientifically valid research purposes, including secondary analyses, meta-analyses, and methodological studies, through the Data Stewards or their designated service providers. Researchers interested in accessing the data should submit a formal request via Population Data BC (www.popdata.bc.ca).

## Acknowledgements

The authors thank the people of British Columbia, whose data are integrated in the administrative health dataset used in this study (BCC19C), and for whom this research is intended to benefit. The authors acknowledge the Data Analytics, Reporting and Evaluation (DARE) team at the Provincial Health Services Authority (PHSA) and BC Centre for Disease Control (BCCDC) for their work in developing, maintaining, and supporting the PHSA’s Platform for Analytics and Data (PANDA).

Conflict of Interest Disclosures: MS has been an investigator on projects funded by GlaxoSmithKline, Merck, Moderna, Pfizer, and Sanofi-Pasteur. DMG has received a speaker honorarium from Roche Diagnostics. All funds have been paid to his institute, and he has not received any personal payments.

Funding/Support: Dr. Watts’s salary was partly funded by the Provincial Health Services Authority through the BC Respiratory Syncytial Virus (RSV) Program. Dr. Viñeta Paramo received a BC Children’s Hospital (BCCH) Research Institute Doctoral Studentship. Dr. Lavoie received salary support from the BCCH Foundation and Research Institute, through the Investigator Grant Award Program. Dr.

Sadarangani is also supported via a salary award from the BC Children’s Hospital Foundation. Dr. Arbour receives salary support through a BCCHR Clinical Investigator Award.

Role of the Funder/Sponsor: The funders had no role in the design and conduct of the study; collection, management, analysis, and interpretation of the data; preparation, review, or approval of the manuscript; and decision to submit the manuscript for publication.

Data Sharing Statement: Access to data provided by the Data Stewards is subject to approval but can be requested for research projects through the Data Stewards or their designated service providers. The following data sets were used in this study: Medical Services Plan Client Roster, Discharge Abstract Database, PharmaNet, Perinatal Data Registry, Sunquest Laboratory Information System, Provincial Laboratory Information Solution and the 2021 Canadian Census. All inferences, opinions, and conclusions drawn in this publication are those of the author(s), and do not reflect the opinions or policies of the Data Steward(s). This Data was provisioned under ISP 21-051. The authors will share the data dictionary upon request.

## SUPPLEMENTARY FIGURES AND TABLES

**Supplementary Figure 1.**
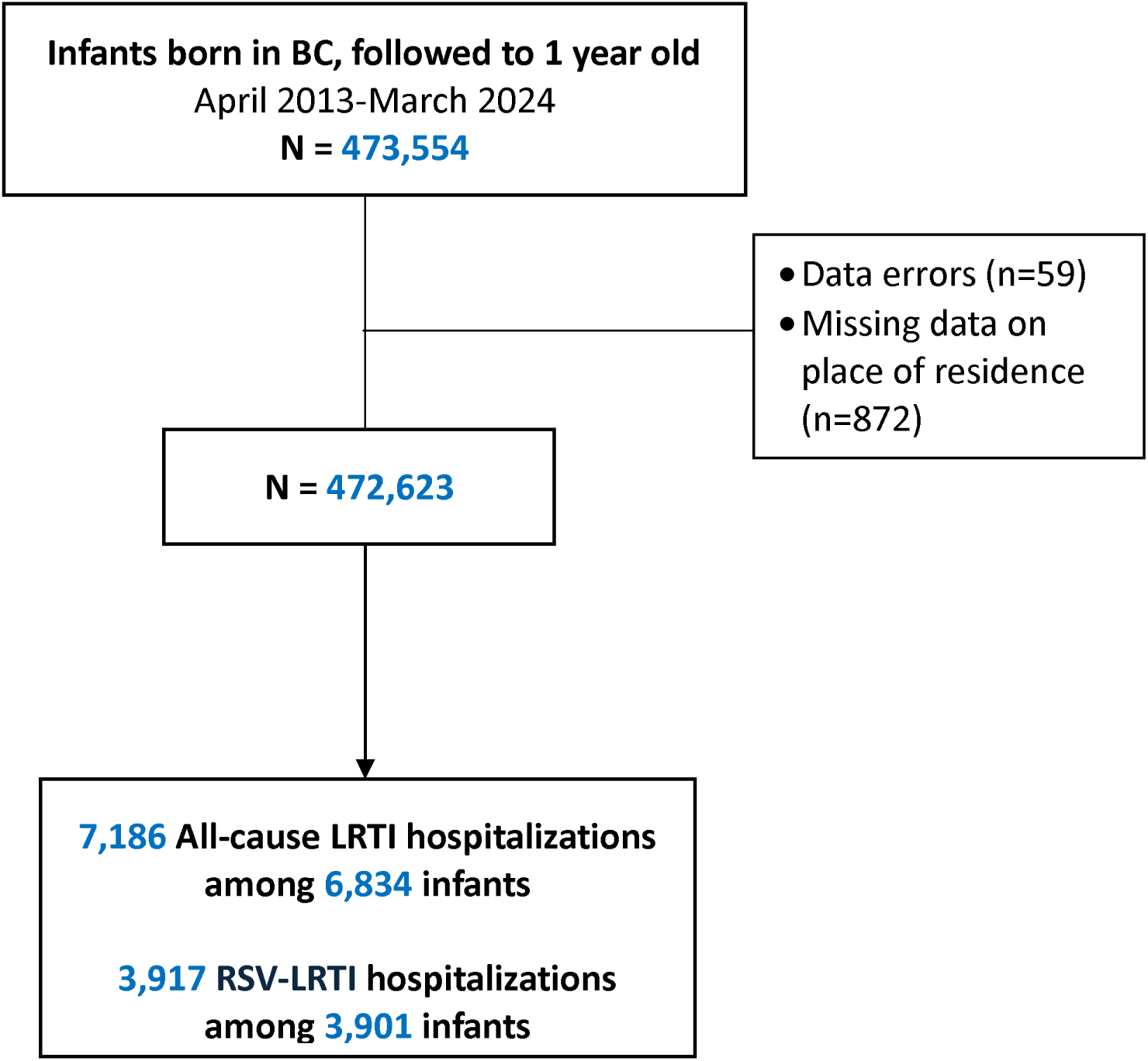
Flow chart, all-cause LRTI and RSV-LRTI hospitalizations in 0–1-year-olds in British Columbia (BC), 2013-2024.

**Supplementary Table 1.** ICD-10 codes used in primary outcome measures.

| **Outcome** | **ICD-10 Code** | **ICD-10 code description** |
| --- | --- | --- |
| RSV-LRTI | J21.0 | Acute bronchiolitis due to RSV |
|  | J12.1 | RSV pneumonia |
|  | J20.5 | Acute bronchitis due to RSV |
|  | B97.4 | RSV as cause of disease classified to other chapters |
| All-cause LRTI | J20-22 | Other LRTIs |
|  | J12-18 | Pneumonia |

**Supplementary Table 2.**
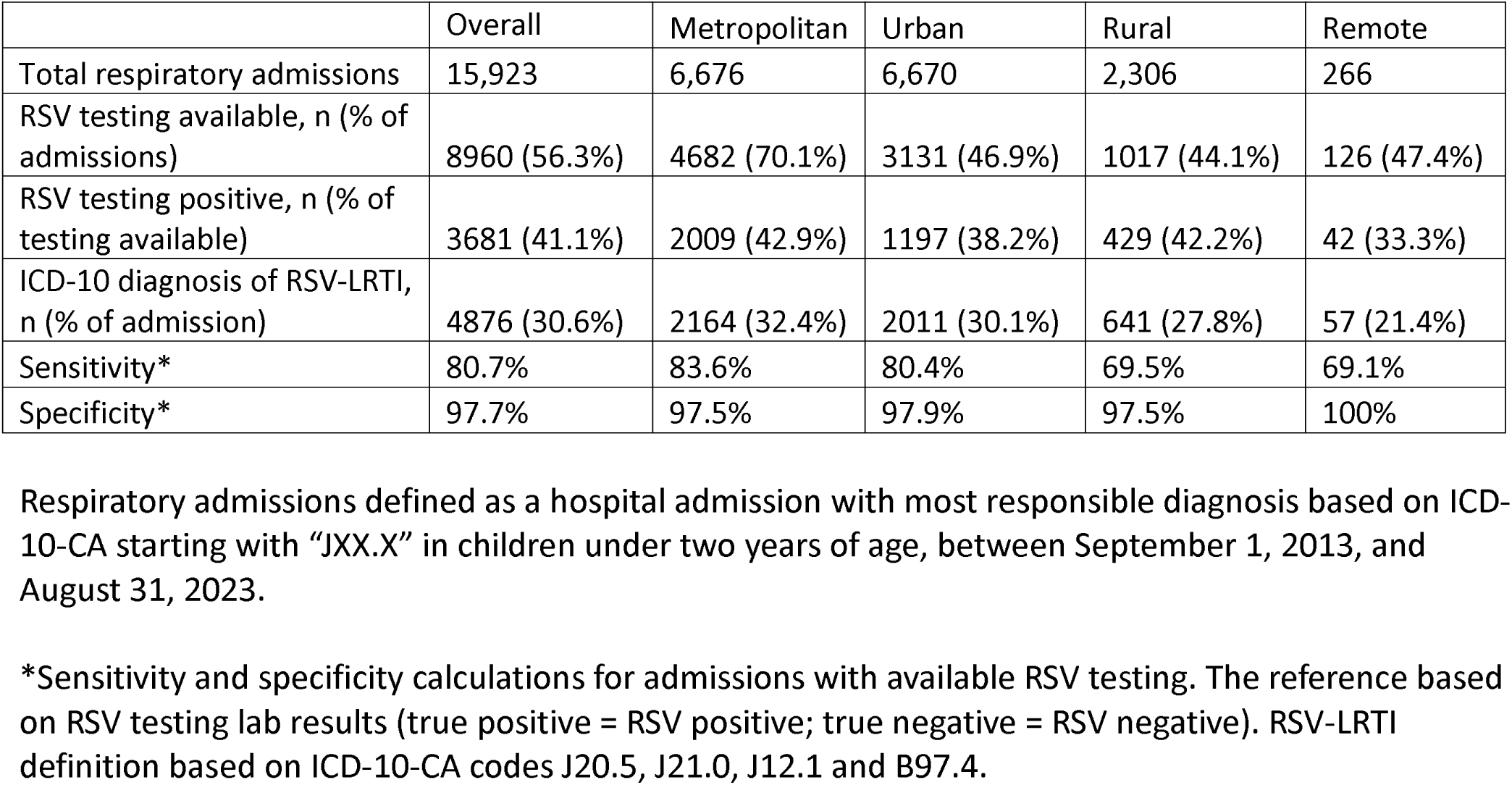
Outcome validation for RSV-LRTI based on ICD-10-CA codes in children under two years of age. Respiratory admissions defined as a hospital admission with most responsible diagnosis based on ICD-10-CA starting with “JXX.X” in children under two years of age, between September 1, 2013, and August 31, 2023. *Sensitivity and specificity calculations for admissions with available RSV testing. The reference based on RSV testing lab results (true positive = RSV positive; true negative = RSV negative). RSV-LRTI definition based on ICD-10-CA codes J20.5, J21.0, J12.1 and B97.4.

**Supplementary Table 3.** Prevalence of chronic medical condition types by organ system involvement among infants by community remoteness (n=472,623). Chronic medical condition sub-categories based on ICD-10 codes at birth based on a modified Pediatric Complex Chronic Conditions Classification^26^, as we described in ^12^.

|  | **Remote**<br>**N=3,636** | **Rural**<br>**N=63,762** | **Urban**<br>**N=164,756** | **Metropolitan**<br>**N=240,469** |
| --- | --- | --- | --- | --- |
| Cardiovascular | 40 (1.1%) | 415 (0.7%) | 1046 (0.6%) | 1551 (0.6%) |
| Congenital or Genetic | 47 (1.3%) | 768 (1.2%) | 2290 (1.4%) | 3439 (1.4%) |
| Endocrine | <5 | 27 (0%) | 64 (0%) | 75 (0%) |
| Gastrointestinal | 14 (0.4%) | 146 (0.2%) | 305 (0.2%) | 450 (0.2%) |
| Hematologic or Immunological | 11 (0.3%) | 95 (0.1%) | 284 (0.2%) | 408 (0.2%) |
| Premature or Neonatal | 92 (2.5%) | 1918 (3%) | 5909 (3.6%) | 9829 (4.1%) |
| Neoplasms | 5 (0.1%) | 31 (0%) | 88 (0.1%) | 134 (0.1%) |
| Neurological | 28 (0.8%) | 281 (0.4%) | 643 (0.4%) | 1241 (0.5%) |
| Renal and Urologic | <5 | 9 (0%) | 18 (0%) | 19 (0%) |
| Respiratory | 14 (0.4%) | 315 (0.5%) | 844 (0.5%) | 1253 (0.5%) |
| Metabolic | 22 (0.6%) | 82 (0.1%) | 212 (0.1%) | 275 (0.1%) |

**Supplementary Table 4.**
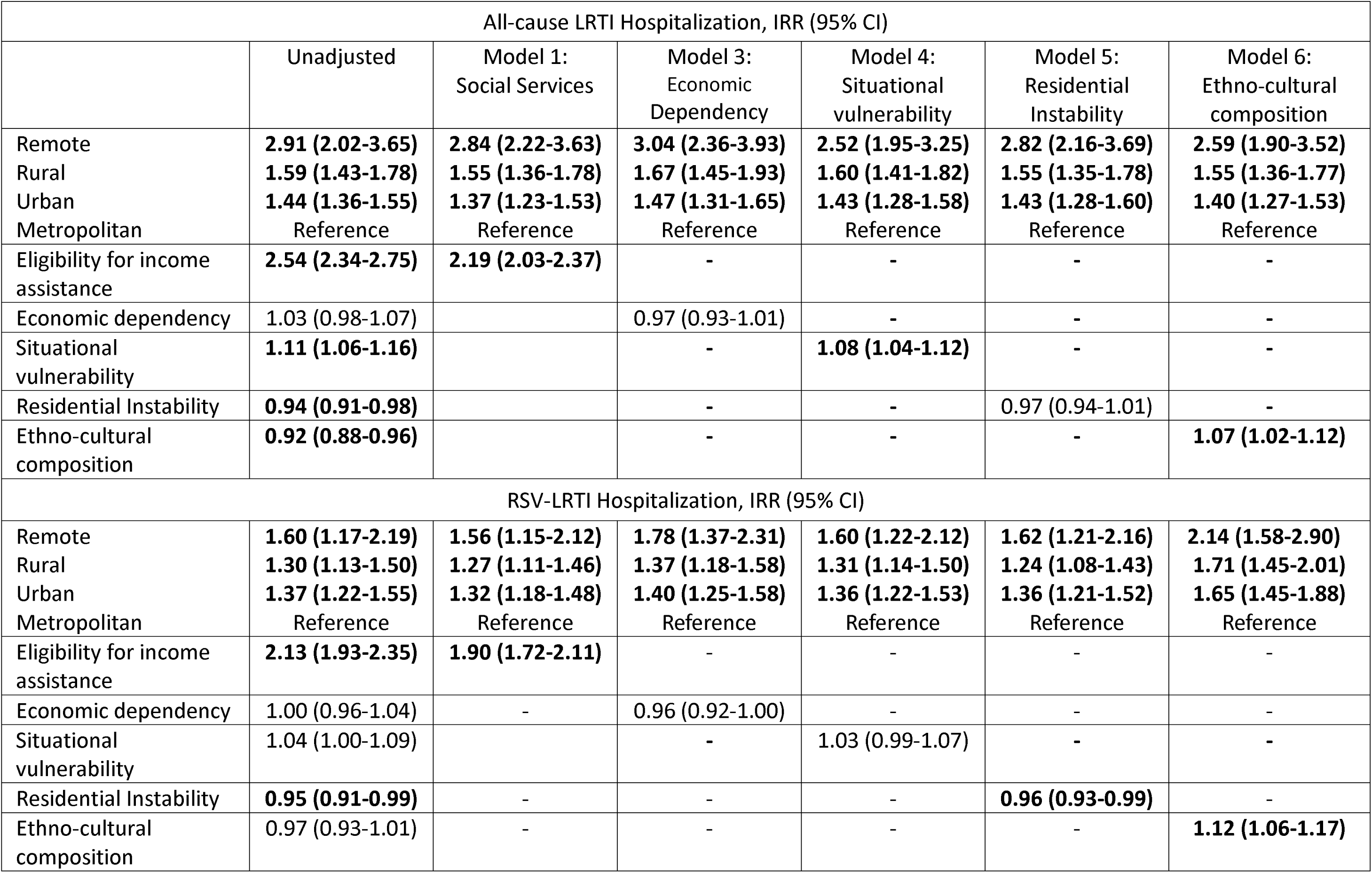
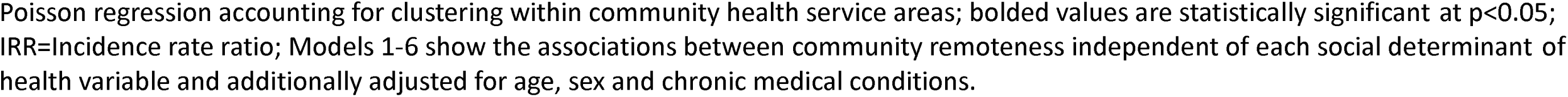
Incidence rate ratios of all-cause and RSV-LRTI hospital admissions by level of community remoteness unadjusted and after adjustment for social determinants of health. Poisson regression accounting for clustering within community health service areas; bolded values are statistically significant at p<0.05; IRR=Incidence rate ratio; Models 1-6 show the associations between community remoteness independent of each social determinant of health variable and additionally adjusted for age, sex and chronic medical conditions.

**Supplementary Table 5.** Incidence rate ratio of all-cause and RSV-LRTI hospitalization for each social determinant of health indicator stratified by community remoteness. Individual Poisson regression models for each social determinant of health indicator adjusted for sex, prematurity and chronic medical conditions and clustering within community health service areas; IRR=Incidence rate ratio; Bolded values are statistically significant at p<0.05. Joint significance test for LRTI (income assistance p=.002, economic=.78, situational=0.04, residential=0.21 and ethno-cultural=0.71) and for RSV-LRTI (income assistance p=.07, economic=.90, situational=0.41, residential=0.15 and ethno-cultural=0.20).

|  | Hospitalization, aIRR (95%CI) |  |
| --- | --- | --- |
|  | All-cause LRTI | RSV-LRTI |
|  | Remote |  |
| Eligibility for Income assistance | 1.73 (0.95-3.12) | 2.32 (0.70-7.68) |
| Economic dependency | 0.95 (0.79-1.13) | 0.90 (0.78-1.03) |
| Situational vulnerability | 2.09 (0.97-4.51) | **1.51 (1.20-1.91)** |
| Residential instability | 1.15 (0.96-1.38) | 1.19 (0.99-1.43) |
| Ethno-cultural composition | 1.08 (0.89-1.30) | 1.01 (0.83-1.24) |
|  | Rural |  |
| Eligibility for Income assistance | **1.61 (1.29-2.00)** | **1.54 (1.22-1.95)** |
| Economic dependency | 0.93 (0.87-1.01) | 0.96 (0.89-1.03) |
| Situational vulnerability | **1.15 (1.06-1.25)** | 1.04 (0.95-1.13) |
| Residential instability | 0.94 (0.83-1.06) | 0.93 (0.83-1.05) |
| Ethno-cultural composition | 1.08 (0.99-1.18) | **1.13 (1.05-1.21)** |
|  | Urban |  |
| Eligibility for Income assistance | **2.21 (1.98-2.46)** | **1.78 (1.55-2.03)** |
| Economic dependency | 0.99 (0.93-1.05) | 0.97 (0.91-1.03) |
| Situational vulnerability | **1.08 (1.03-1.14)** | 1.03 (0.97-1.11) |
| Residential instability | 0.99 (0.94-1.06) | 0.99 (0.94-1.05) |
| Ethno-cultural composition | 1.06 (0.99-1.15) | **1.15 (1.07-1.24)** |
|  | Metropolitan |  |
| Eligibility for Income assistance | **2.59 (2.32-2.89)** | **2.31 (1.96-2.71)** |
| Economic dependency | 0.97 (0.89-1.04) | 0.95 (0.88-1.03) |
| Situational vulnerability | 1.04 (0.99-1.10) | 1.01 (0.95-1.08) |
| Residential instability | **0.95 (0.91-0.99)** | **0.94 (0.90-0.99)** |
| Ethno-cultural composition | 1.03 (0.92-1.15) | 1.02 (0.90-1.13) |

